# Contextualizing covid-19 spread: a county level analysis, urban versus rural, and implications for preparing for the next wave

**DOI:** 10.1101/2020.04.24.20078204

**Authors:** J. Sunil Rao, Hang Zhang, Alejandro Mantero

**Affiliations:** Division of Biostatistics, University of Miami

**Author notes:** indicates corresponding author.

**Keywords:** Covid-19, Pandemic, Contextual Factors, County Level, Growth Curves, Random Forests, Machine Learning

## Abstract

Paraphrasing [Morano and Holt, 2017], contextual determinants of health including social, environmental, healthcare and others, are a so-called deck of cards one is dealt. The ability to modify health outcomes varies then based upon how one’s hand is played. It is thus of great interest to understand how these determinants associate with the emerging pandemic covid-19. To this end, we conducted a deep-dive analysis into this problem using a recently curated public dataset on covid-19 that connects infection spread over time to a rich collection of contextual determinants for all counties of the U.S and Washington, D.C. Using random forest machine learning methodology, we identified a relevant constellation of contextual factors of disease spread which manifest differently for urban and rural counties. The findings also have clear implications for better preparing for the next wave of disease.

## 1 Introduction

Like politics, all public health is ultimately local. This tenet is particularly important in trying to understand the dynamics of an emerging outbreak like the pandemic covid-19. While much attention has focused on modeling infection spread and containment and mitigation strategies to better plan for potential surges and the impact they may have on hospital capacities, less attention has been paid to understanding the contextual nature of the pandemic.

This means modeling area level data like at the county level and including information on demographics, race/ethnicity, housing, education, employment and income, climate, transit scores and healthcare metrics [Killeen et al., 2020]. Many of these are often classified under the umbrella of social determinants of health that have been documented to influence the success or setbacks in steps to a successful public health emergency response [Morano and Holt, 2017]. Being able to do this at a local level like the county has significant implications for understanding how where you live influences disease spread, differences between rural and urban settings, and better planning for future anticipated waves of disease. In the absence of comprehensive geocoded patient level data where multilevel interactions can be assessed, the study of the area level influences alone on covid-19 spread is informative in its own right. In addition, with the realization that racial and ethnic minority groups are suffering disproportionately in the pandemic, social determinants of health (which are important contributors to health disparities) have garnered more attention.

To that end, recently [Killeen et al., 2020] designed a dataset containing a machine-readable file with demographic, socioeconomic, healthcare and education data for each county in the 50 states and Washington, D.C. organized by FIPS codes which are unique county code identifiers. This data was cobbled together from 10 governmental and academic sources on the county level. The resulting dataset contains more than 300 contextual county level variables. They then merged this contextual data with county-level time series data from the JHS CSSE covid-19 dashboard [Dong et al., 2020] which gives continuously updated confirmed infection and death counts over time.

Our goal in this paper is to estimate growth rate increases (slopes) of the covid-19 disease for each county in the U.S. and then relate these estimates to contextual level factors. In addition, we would like to understand differences between urban and rural county (i.e. populations < 50,000) settings in terms of the contextual factors that most relate to growth rates. Since we expect these factors to interact in complex ways, we will employ random forests machine learning methodology [Breiman, 2001] for model fitting. We expect that our findings will help to inform more nuanced strategies for safely re-opening society at the local level.

## 2 Methods

### 2.1 Study population

[Killeen et al., 2020] assembled the dataset under focus. Covid-19 infection volume time series came from the Johns Hopkins University CSSE COVID-19 Tracking Project and Dashboard which when the data was pulled ranged from 01/22/2020 until 04/12/2020. Climate data came from NOAA. County level demographic data was extracted from the US Census Bureau and the USDA. Healthcare capacity related data came from the Kaiser Family Foundation. Traffic score information came from the Center for Neighborhood Technology. ICU bed information came from Kaiser Health Network. Additional covid-19 case information came from USAfacts. Physcian workforce data came from AAMC.

Many of the variables were not usable in their original forms, many involved counts within a county but did not take into account the overall size of the county itself. This would make it impossible to directly compare between counties. Therefore, all count variables were standardized by their totals for example counts of racial and gender groups were divided by the total population of the county and household counts were divided by the total number of households in the county. These variables are listed in the list of columns section of the GitHub repository created to provide this data [Killeen et al., 2020].

There was also an issue of state level variables that were included in the dataset either by copying the state level data for every corresponding county or by extrapolating the state characteristic as a fraction of population. Both types of variables were excluded from the analysis. Our reasoning for this is that the state level information could muddle the mechanism for the individual counties since our goal is to discern the slopes specifically at this level. If the forest were to start grouping counties by data it would defeat the purpose of the analysis. As for the extrapolated values by population, this is a very strong assumption that will most likely overstate the resources available to rural counties. Furthermore, this again may cause the forest to group by state which we want to avoid, our point is to have it group by specifically county level characteristics thus giving us groups of similar counties that experience similar increases in infection rates.

Highly correlated variables were removed because they represented very similar information. The vast majority of these removed variables were upper and lower bound estimates of means included in the dataset. There is one pair of variables for migration that we suspected would be highly correlated, one was absolute count and the other was rate, for these the correlation was not very high and we though both could be particularly important in predicting slopes so both were included.

We also removed any variables with a missingness rate of over 45%. This resulted in a final set of 186 variables that were analyzed. Full codebook for variable descriptions on Github site accompanying [Killeen et al., 2020]. Missing values in these were handled using random forest imputation (discussed in the next subsection).

Some counties had to be omitted from the analysis and this was mostly due to poor fit in the `growthcurver` procedure. Either the model was not estimable or the estimated midpoint time was much farther out than the data that was available at the time of analysis. We wanted to avoid such far extrapolations when the actual data only existed for far fewer days. This resulted in a loss of 16.8% of the counties. Also, for any counties that continued to have zero infections although the model could not be fit, we assumed the slopes to be zero.

### 2.2 County-specific confirmed infections growth curve fitting and growth rate estimation

The infections time series county-level data was obtained from [Killeen et al., 2020] that updated on Apr 13, 2020. The counts of infection in each county were standardized by corresponding county population and then fit into growth curve model. The growth curves for each county was estimated based on logistic equation which is commonly used in population ecology [Crow et al., 1970; Rockwood, 2015; Sprouffske and Wagner, 2016] utilizing R package `growthcurver` [sprouffske, 2018]:

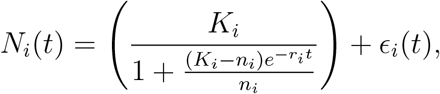

where *N*_*i*_(*t*) represents the proportion of infected people at time t for a given county *i, n*_*i*_ as the size of initial infection proportion, *K*_*i*_ as the carrying capacity, *r*_*i*_ is the growth rate with respect to the infection proportion, and *ϵ* _*i*_(*t*) the model errors assumed to have zero mean and constant variance. Non-linear least squares is used to estimate model parameters.

The growth curve slopes were further obtained by taking derivative of *N*_*i*_(*t*) with respect to *t* for each county as:

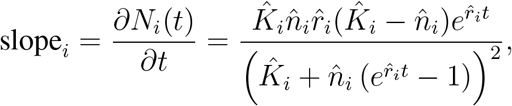

where estimated parameter values are indicated by a hat sign. Then to calculate the maximal slope, we plug in the midpoint of time for *t* which for this type of function represents when the change in the outcome is greatest.

### 2.3 Random Forests regression

Random forest (RF) is a machine learning technique developed by Leo Brieman [Breiman, 2001] which has been implemented into many subject matters. The technique takes advantage of base learner aggregation to develop a complex learner that has much improved prediction performance and fit on the data. In this case the base learners are CART trees [Breiman et al., 1984] but in order to prevent having the same tree appear multiple times in the forest, randomness is introduced to the learning procedure. This takes the form of both bootstrapping the observations for the training of each tree and random feature selection where the optimal split is not taken from all variables available but instead a randomly selected subset. This results in the trees being more independent of each other thus preventing the instability that is present in CART. The issue with CART tree is the balancing between bias and instability via tree depth, if the tree grows too deep, then it is likely have overfit the data (high instability) but will have low bias; while, if it grows too shallow, it will be more stable but have higher bias. Therefore by aggregating very deep and unstable trees random forest is able to obtain overall low bias and average away the instability of each tree thus giving us a very effective overall learner.

We used the `rfsrc` function in R [Ishwaran and Kogalur, 2020] for fitting random forests. Growth curve slope estimates were treated as responses (*y*) and the 186 contextual variables described above as county level predictors (*x*_*j*_; *j* = 1, …, 186). We chose not to transform the responses because of the robustness of random forests and to facilitate model interpretations ([Shah et al., 2014]). Random forest procedures default settings were utilized which included `mtry` of square root of p (i.e. the number of predictors). Additionally, the automatic imputation function that randomForestSRC [Ishwaran and Kogalur, 2020] was activated in order to avoid losing county information. The imputation procedure is built into the forest growing procedure and run all at once. We included imputation because if we were to run the model only on the complete data, we would have lost approximately one third of the counties with the biggest loss being in the rural counties. In order to avoid losing so much data, we opted for imputation in our RF training. Separate forests were fit to rural and urban counties. Relative variable importance plots were generated and models validated using the % variance explained defined as,

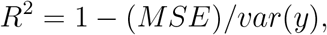

where *MSϵ* is defined as the out-of-bag test error estimated on observations not showing up in bootstrap resamples. Values near zero (or negative for that fact) are indicative of modeling noise [Breiman, 2001]. Relative variable importance is obtained by dividing all the calculated importances by the maximal one, this then provides a value with a possible range between 0 and 1 which makes it easier to interpret. Values near zero (or negative for that fact) are indicative of modeling noise [Breiman, 2001]. Visualization of RF fits were done using parallel coordinate plots and multi-variable co-plots. These are created based on clusters which were determined with the PAM method using random forest distances as described in [Mantero, 2018]. For PAM, *k* was set to 10, the goal here was not to obtain the optimal number of clusters but to separate out a few groups with differing average slopes in order to depict how the contextual variables change as the slopes increase or decrease by group.

Random forest distances work similarly to random forest proximity but are more sensitive to the tree topology therefore able to provide better clusters. Like random forest proximity, a pair of points are considered highly proximal if they are in the same terminal node, but unlike proximity, instead of simply assigning zero if they are not in the same terminal node, a number between 0 and 1 is used depending on how far down the tree the two points became disjoint.

In order to attempt to ascertain potential differences in how rates of infection increases relate to the contextual variables between urban and rural settings, rural county data was dropped down the RF trained on urban data and urban county data was dropped down the RF trained on rural data. For this type of reverse prediction, the resulting errors were calculated and 95% confidence intervals determined. This allow us to see if there were regions of non-overlap suggesting the presence of possible heterogeneity between the associations between the rates with their contextual variables depending on rural or urban settings.

## 3 Results

Figure 1 shows sample growth curves fit to two large urban counties (New York, NY(population 1628701) and Miami-Dade, FL(population 271581)) and two rural counties (Randolph county, GA (population 6833) and Barbour county, AL (population 24881)). New York county and Randolph county had large slope estimates and Miami-Dade and Barbour counties had much smaller slopes estimates (note the much smaller values of the y-axis for Babour county). Figure 2 shows the estimates growth curve slopes versus log_10_(population) based on 2019 census population values or all counties in the U.S. Blue coloring indicates rural counties and red coloring urban. While the majority of estimated slopes are not large, it is clear that there are a mixture of urban and rural counties with large slope estimates.

**Figure 1:**
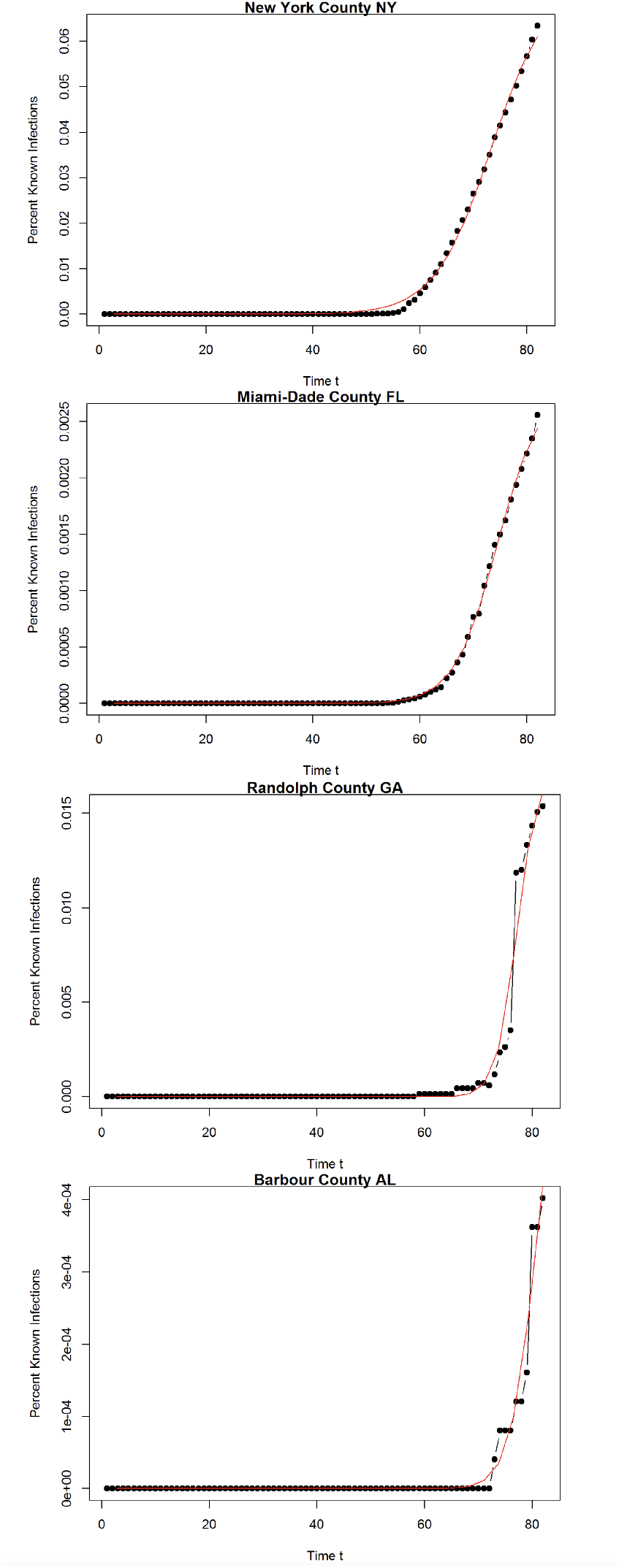
A sample of two urban and two rural counties - one in each group with a large slope estimate and the other with a smaller slope estimate.

**Figure 2:**
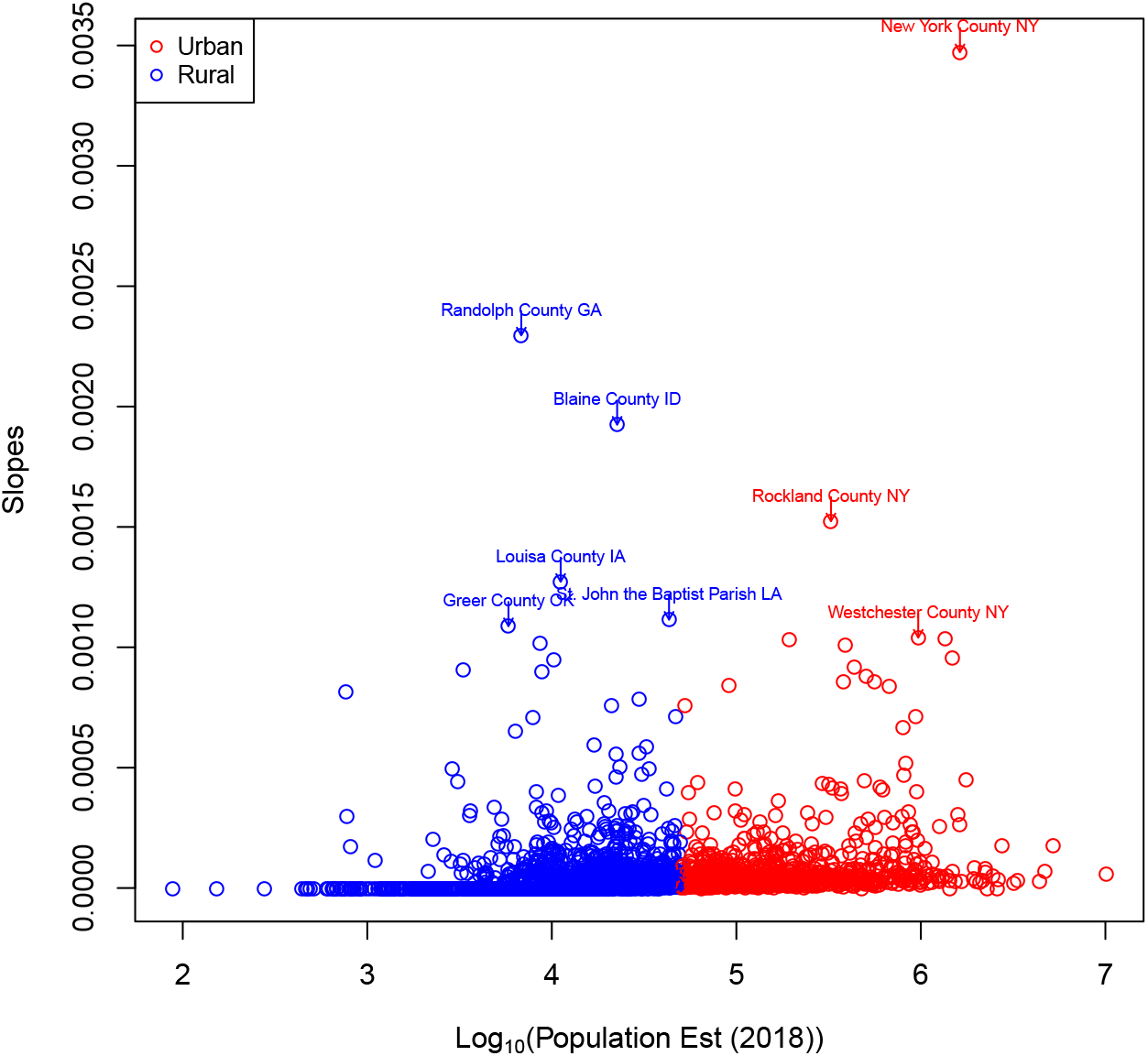
County-specific growth rate estimates (slopes) versus *log*_10_(*population*) based on 2018 census numbers. Coloring by rural or urban status.

Random forests were fit separately to urban and rural counties. For the urban forest the % variance explained was 32.26% and 32.56% for the rural forest. This is very much on par with other studies that focus on area-level variables only (see for example, [Richardson et al., 2017], [Oliveira et al., 2012]). This shows good model validation for both forests.

Tables 1 and 2 display relative variable importance tables (i.e. mean decrease in accuracy using out-of-bag prediction when the variable is permuted) for urban counties and rural counties respectively. These were extracted from the relative variable importance scree plots in Figure 3 by choosing those variables with large relative importance values left of the visible “elbow” in the plots.

**Table 1:** Important variables for urban RF

| Predictors | Description | Rel. importance |
| --- | --- | --- |
| HBA_FEMALE | Hispanic, Black or African American alone female population | 1.000 |
| HBA_MALE | Hispanic, Black or African American alone male population | 0.978 |
| NHIAC_MALE | Not Hispanic, American Indian and Alaska Native alone or in combination male population | 0.949 |
| HBAC_FEMALE | Hispanic, Black or African American alone or in combination female population | 0.785 |
| NHIA_MALE | Not Hispanic, American Indian and Alaska Native alone male population | 0.775 |
| INTERNATIONAL_MIG_2018 | Net international migration in period 7/1/2017 to 6/30/2018 | 0.684 |
| Density.per.square.mile.of.land.area...Housing.units | Land area density for housing units as per 2010 census | 0.655 |
| Density.per.square.mile.of.land.area...Population | Land area density for population as per 2010 census | 0.562 |
| May.Temp.AVG...F | Average temperature in May, 2019 in Fahrenheit | 0.449 |
| May.Temp.Max...F | Maximum temperature in May, 2019 in Fahrenheit | 0.422 |
| HBAC_MALE | Hispanic, Black or African American alone or in combination male population | 0.378 |
| NHTOM_FEMALE | Not Hispanic, Two or More Races female population | 0.372 |
| Apr.Temp.Max...F | Maximum temperature in Apr, 2019 in Fahrenheit | 0.335 |
| R_INTERNATIONAL_MIG_2018 | Net international migration rate in period 7/1/2017 to 6/30/2018 | 0.312 |
| NHTOM_MALE | Not Hispanic, Two or More Races male population | 0.297 |
| NHIA_FEMALE | Not Hispanic, American Indian and Alaska Native alone female population | 0.282 |
| May.Temp.Min...F | Minimum temperature in May, 2019 in Fahrenheit | 0.273 |
| Jun.Temp.Min...F | Minimum temperature in Jun, 2019 in Fahrenheit | 0.272 |
| DOMESTIC_MIG_2018 | Net domestic migration in period 7/1/2017 to 6/30/2018 | 0.268 |
| NHNAC_FEMALE | Not Hispanic, Native Hawaiian and Other Pacific Islander alone or in combination female population | 0.235 |
| Total.households..Nonfamily.households.. Householder.living.alone 65.years.and.over | Total number of nonfamily households where Householder living alone is 65 years and over | 0.232 |
| MEDHHINC_2018 | Estimate of median household income 2018 | 0.211 |
| NHIAC_FEMALE | Not Hispanic, American Indian and Alaska Native alone or in combination female population | 0.194 |
| Apr.Temp.AVG...F | Average temperature in Apr, 2019 in Fahrenheit | 0.192 |
| NHNA_FEMALE | Not Hispanic, Native Hawaiian and Other Pacific Islander alone female population | 0.189 |

**Table 2:**
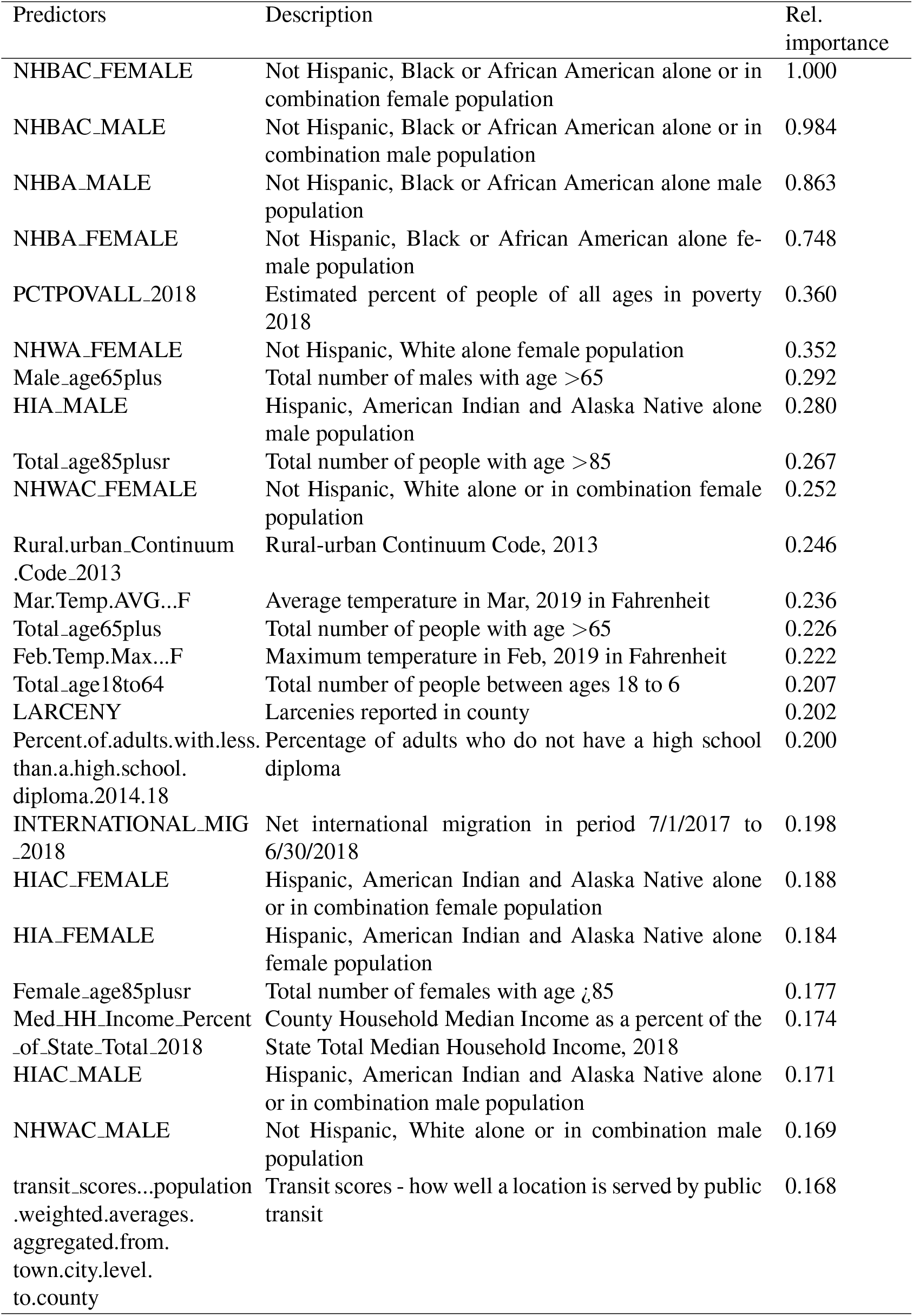
Important variables for rural RF

**Figure 3:**
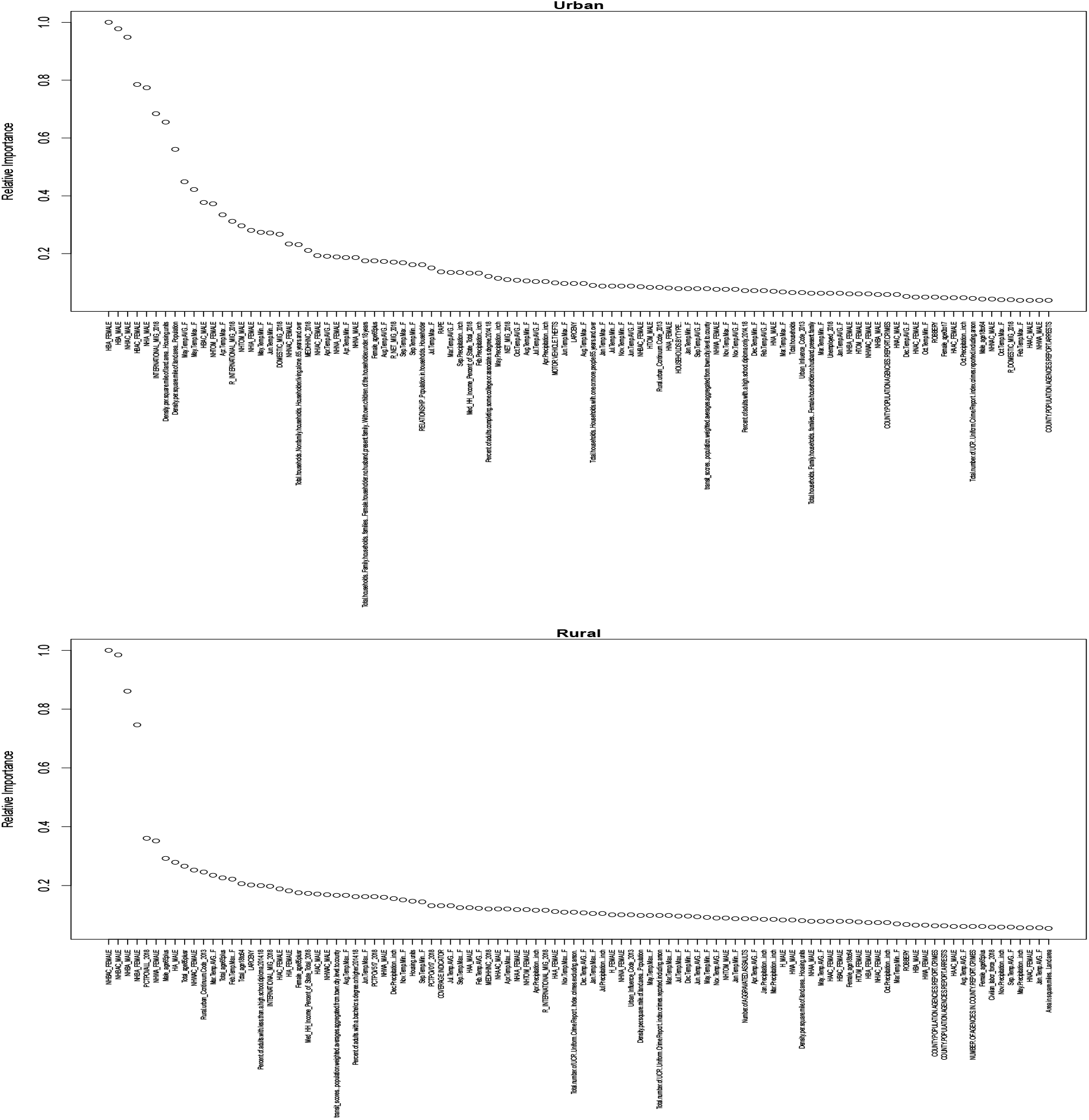
Variable relative importance scree plots: urban (top), rural (bottom).

In order to better visualize the fitted random forests, we next categorized counties by slope estimate groupings as determined by the fitted forests. Parallel coordinate plots are shown in Figure 4 and Figure 5 for urban and rural counties respectively. Each group is represented by lines moving horizontally and each variable by a vertical line (whose range of values is above and below the vertical line). Lines connect mean values of each variable for that group. The mean value of the slope for the group is shown in the first vertical line. The variables are sorted according to relative variable importance.

**Figure 4:**
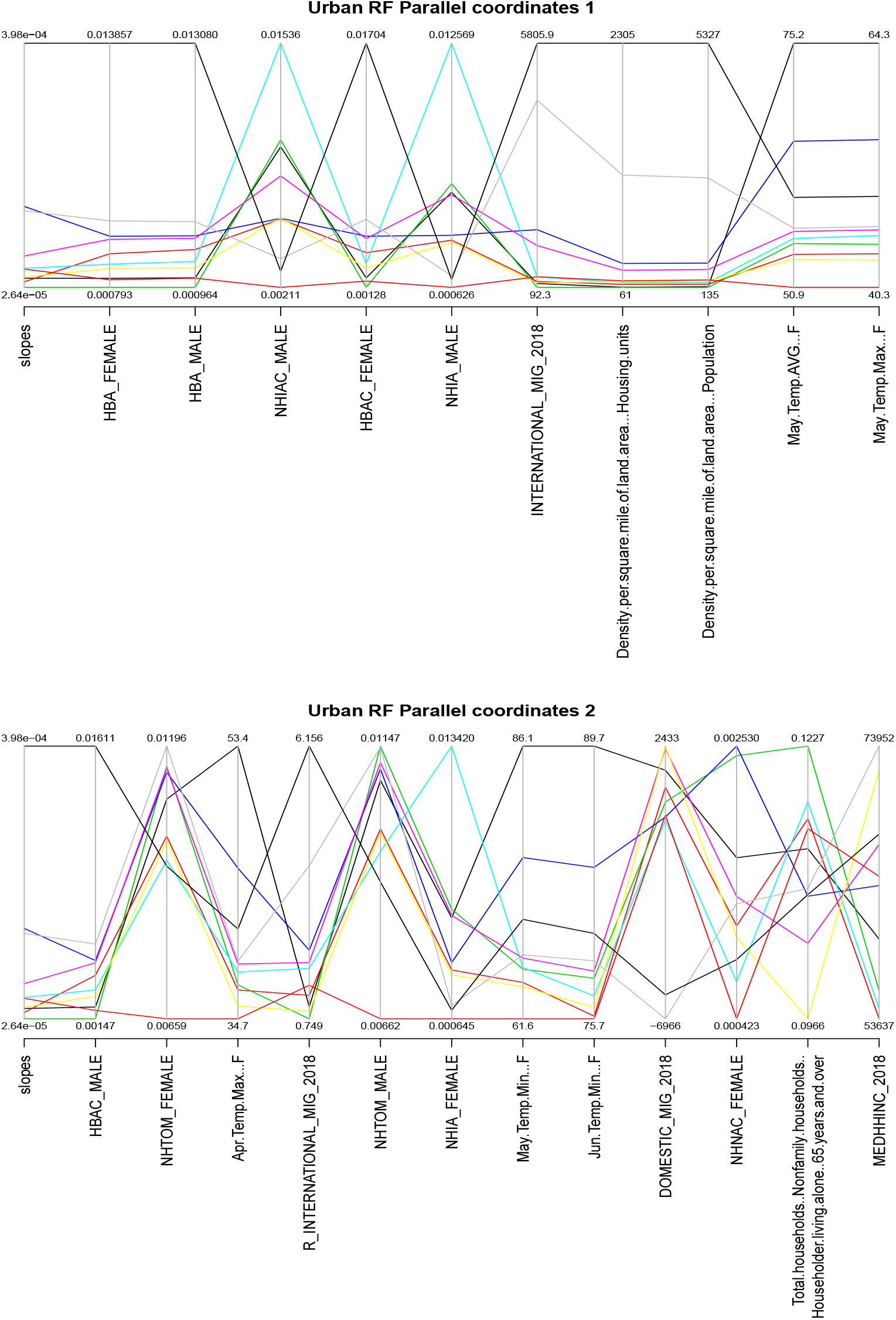
Parallel coordinate plots for urban counties.

**Figure 5:**
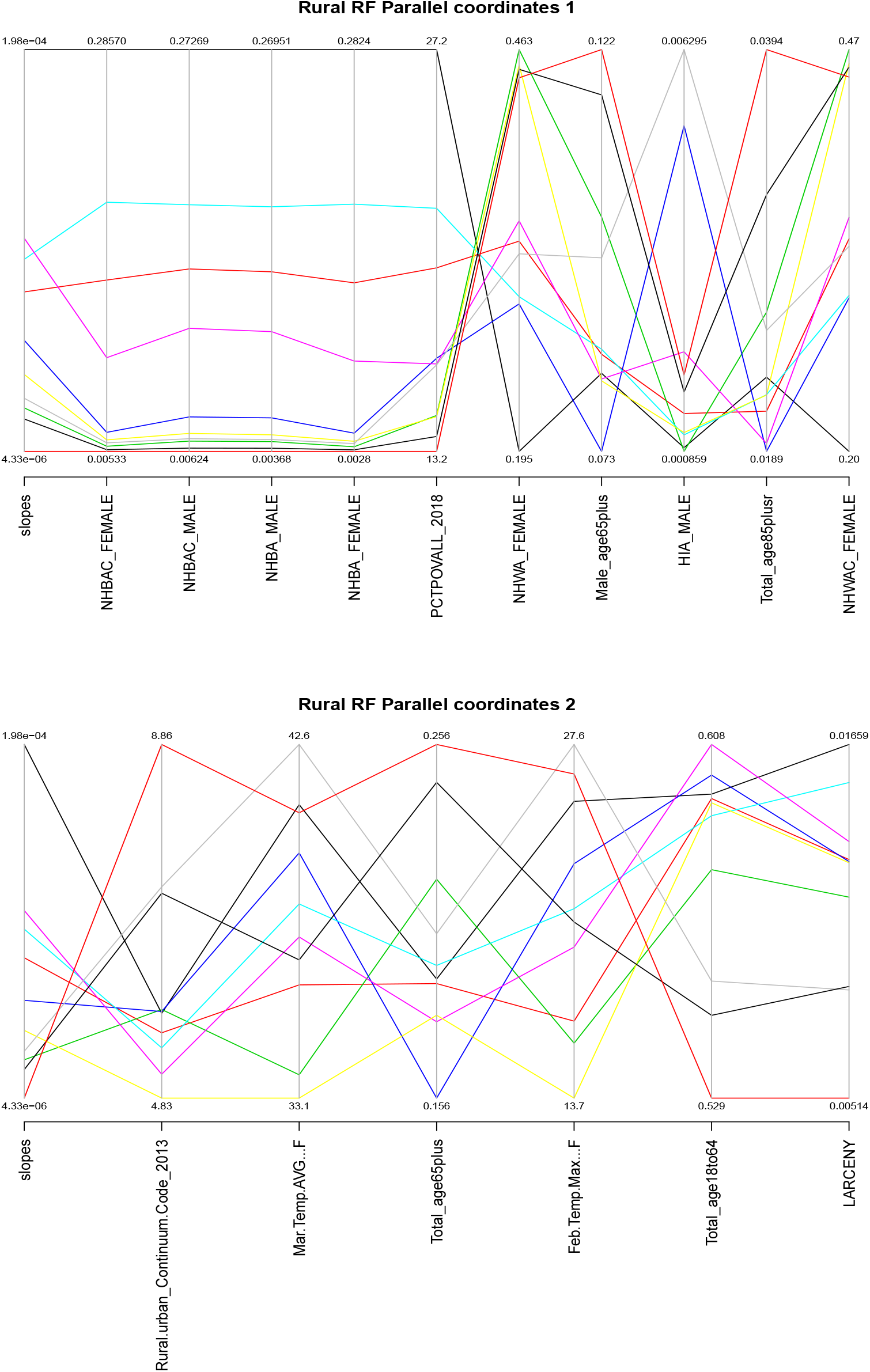
Parallel coordinate plots for rural counties.

It’s clear that there are clusters of variables that clearly separate slope groups and that the variables differ for the urban versus rural random forest. For the urban parallel coordinate plot, such variables included percentage of Hispanic, black or African American male and female (HBA_FEMALE, HBA_MALE), international migration (INTERNATIONAL_MIG_2018), housing density, population density and May 2018 average and maximum temperature values in degrees F. For rural counties, such variables included percentage of non-Hispanic, black or African American male and female (NHBAC_FEMALE, NHBAC_MALE, NHBA_MALE, NHBA_FEMALE), 2018 and percent poverty as estimated in 2018. Other variables of rural importance also included age - specifically the percent of the population that were elderly.

Figure 6 and Figure 7 show co-plots focusing on these key variables. This allows a closer inspection of how the growth curve slope estimates continuously change as a function of other variables. In particular each co-plot showed codependence on 4 other variables. For the urban co-plot, variables plotted include HBA_MALE, population density, and 2018 international migration numbers. Each line is shown with a different color and plotting character. As the legend indicates, they represent quintiles of the 2018 average May temperature. There are some clearly interesting patterns where slopes are seen to rise dramatically. For instance, the top right panel indicates that for counties with higher average 2018 May temperatures, higher percent black male populations and higher degrees of international migration, that covid-19 spread much more dramatically than in other urban counties.

**Figure 6:**
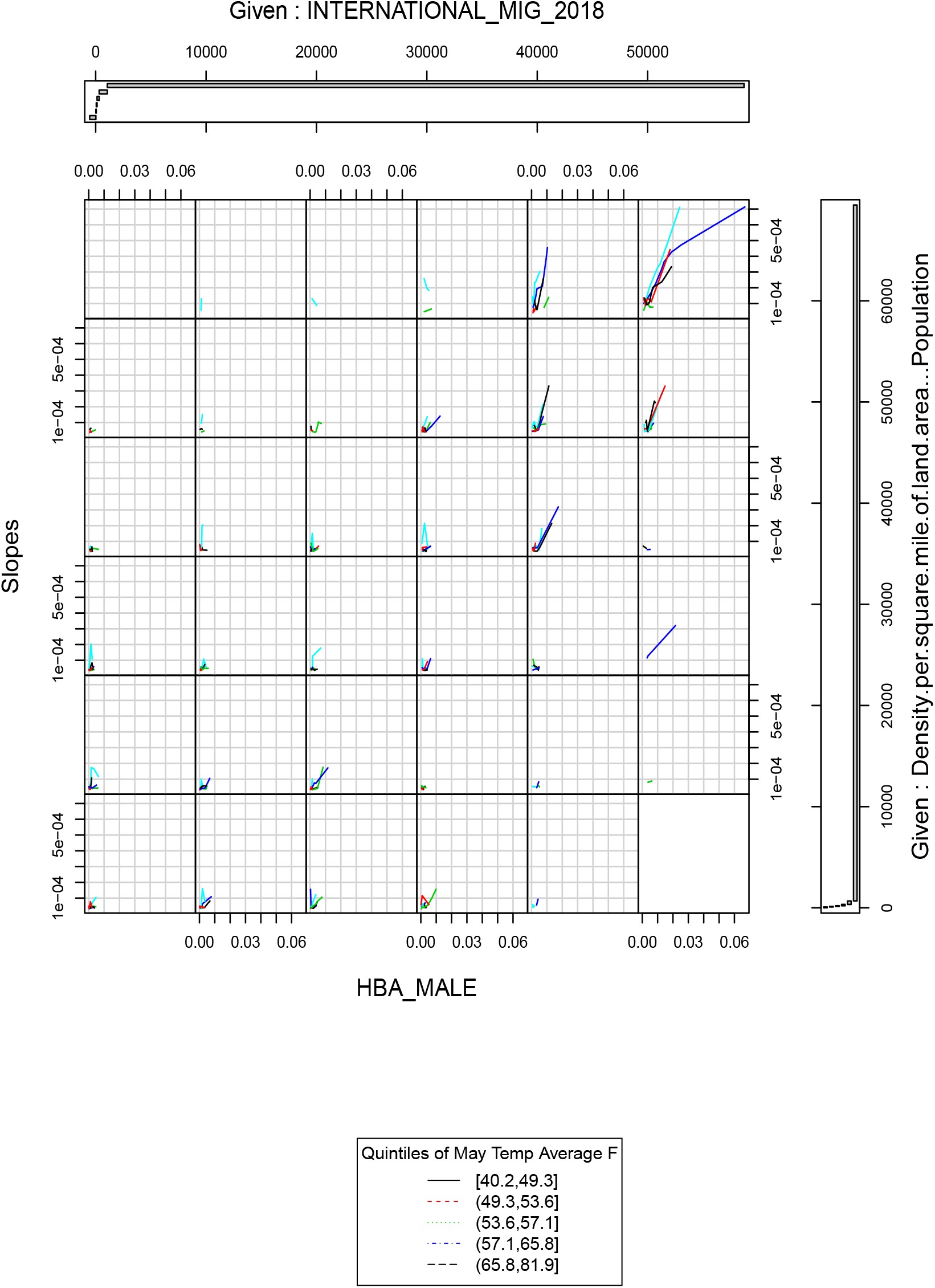
Co-plot for urban counties.

**Figure 7:**
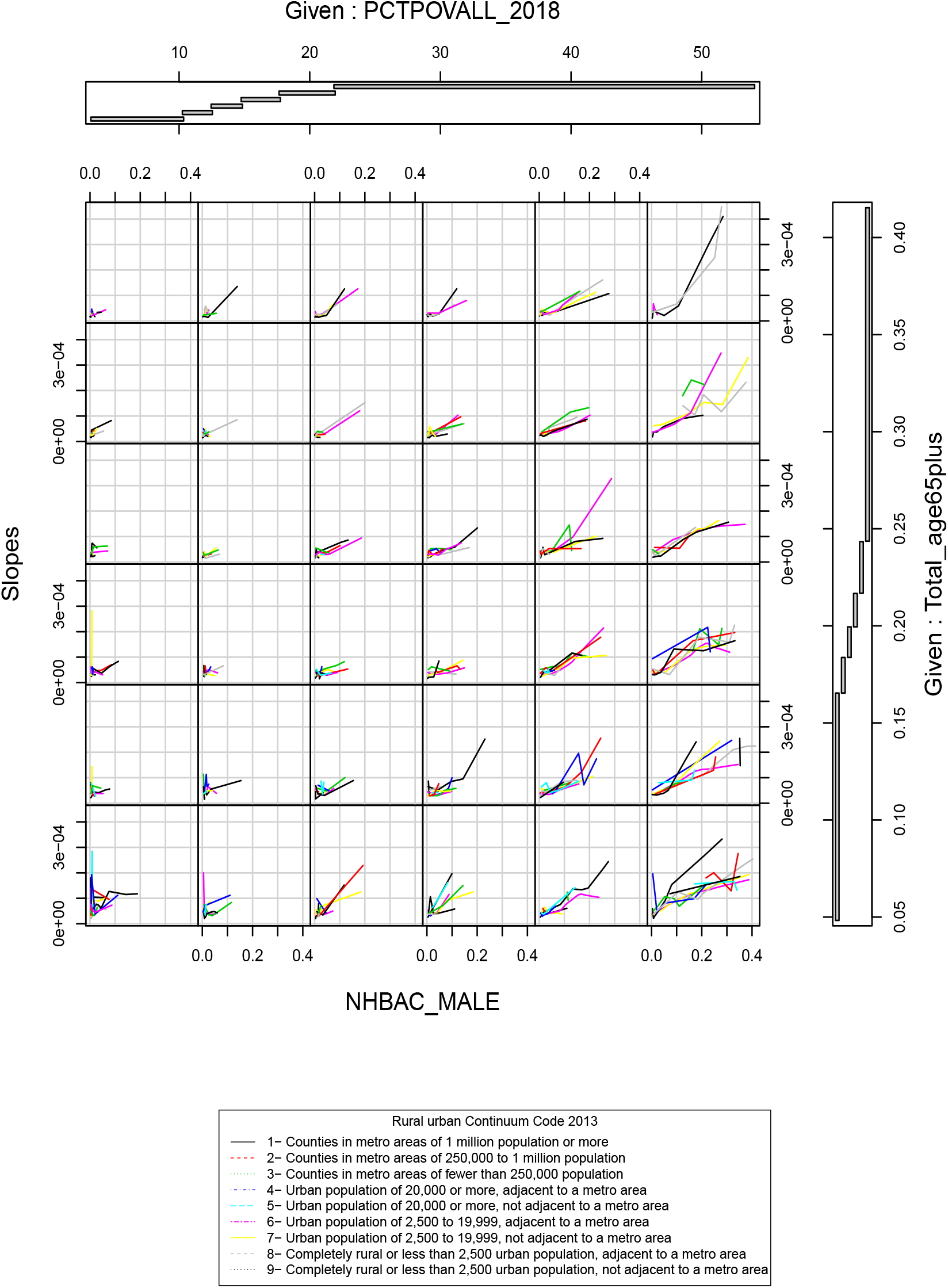
Co-plot for rural counties.

Contrast this with the rural co-plot. There are multiple interesting panels here but focus again on the top right panel. Here each line represents a group of counties classified by rural-urban continuum codes in 2013 with larger numbers indicating smaller population counties either adjacent or not adjacent to a metro area. Smaller, (and also more isolated) counties saw a much more rapid increase in covid-19 spread when accompanied by having a proportionately more black, older population who also had a higher percentage of their population in 2018 living in poverty.

Reverse predictions for urban counties were generated from the rural forest and for rural counties from the urban forest. Absolute error 95% confidence intervals were [4.967 × 10^−5^, 5.513× 10^−5^] and [7.224 ×10^−5^, 8.535 ×10^−5^] respectively. The non-overlap further confirms some differences in county level structure being captured by both forests.

## 4 Discussion

Social determinants of health are known to influence the spread of infectious diseases [Bishwajit et al., 2014]. These include the impact of poverty, illiteracy, poor sanitation, food insecurity, overcrowding, access to healthcare, workplace conditions and transit options amongst others. Other important factors are immigration patterns, age distributions, weather and environmental conditions. These factors can play a large role in determining how successful the response to a public health emergency is [Morano and Holt, 2017]. They also are intimately involved in health disparities that arise in more acute fashion during a pandemic. This is already been observed for the covid-19 pandemic where people of color have borne a disproportionate burden of mortality and morbidity of the disease.

Abiding by the tenet that public health is ultimately local in nature, we focused on modeling county level data about disease spread and county-level determinants. By linking confirmed infection numbers via growth curve modeling to a large number of area-level factors, we used random forest modeling to show interactions between area-level factors that associate with increased disease spread. These associations were quite different for urban versus rural counties. Urban county spread was more influenced factors like racial distributions (in particular the concentration of African American individuals in a county), population and housing density, international immigration patterns and cyclical weather patterns. Rural county spread was also influenced by similar racial composition, but also by poverty, the degree of elderliness and the degree of rurality.

The random forest models demonstrated objective goodness-of-fit on par other studies that focused on area-level predictors alone. These models can be particularly good at identifying complex interactions and so were well suited for this analysis. As noted, random forest imputation was performed and so we did a sensitivity analysis to confirm that the results were not overly influenced by the imputation scheme [Resseguier et al., 2011].

There are some ways in which this analysis could be improved. First, we did not take into account particular county level interventions that were made over time - for example lockdowns. This data does in fact exist as another data file in the same repository at Johns Hopkins University. One strategy to incorporate this data would be to add in lockdown status as a time-varying covariate in the growth curve model and then re-estimate growth curve slopes. One strategy for this is to use group counties by lockdown dates and then use the technology of non-linear mixed models using additional covariates [Pinheiro, 2002].

Further information that would have been useful to include in the model but was not available at the county level includes number of nursing home facilities and traffic flow or movement data. The nursing home facilities although somewhat presented in the age demographics of the county, are important in their own right since they often served as sites of localized case clusters. Traffic flow or movement data would indicate the degree to which lockdown measures were being adhered to. Lastly, one missing confounding variable is the uneven use of testing across different regions of the country. This would clearly affect the confirmed infection count time series numbers.

## Data Availability

The data is all publicly available.

https://github.com/JieYingWu/COVID-19_US_County-level_Summaries

## Acknowledgements

JSR was partially supported by NSF grant DMS 1915976 and NIH grants U54 MD010722 and UL1 TR000460. HZ and AM were partially supported by NIH grant UL1 TR000460. The authors would like to thank Dr. Kevin Patel at the University of Washington for very helpful discussions.

## Notes

### Competing Interest Statement

The authors have declared no competing interest.

